# Examining a Preclinical Alzheimer’s Cognitive Composite for Telehealth Administration, the tPACC, for Reliability between In-Person and Remote Cognitive Testing with Neuroimaging Biomarkers

**DOI:** 10.1101/2023.08.03.23293329

**Authors:** Tugce Duran, Sarah A. Gaussoin, Lauren A. Latham, Melissa M. Rundle, Mark A. Espeland, Benjamin J. Williams, Timothy M. Hughes, Suzanne Craft, Bonnie C. Sachs, James R. Bateman, Samuel N. Lockhart

## Abstract

**INTRODUCTION:** We defined a telehealth preclinical Alzheimer’s cognitive composite (tPACC) derived from remotely administered cognitive assessments and examined its relationship with neuroimaging biomarkers.

**METHODS:** We examined neuroimaging and in-person and remote cognitive testing data from the Wake Forest Alzheimer’s Disease Research Center’s Clinical Core cohort and a pilot study to form a modified PACC (PACC5-RAVLT) and tPACC. We performed intraclass correlation coefficient (ICC) analysis for reliability of tPACC and linear regression models to evaluate associations between tPACC and neuroimaging. Bland-Altman plots for agreement were constructed across cognitively normal (NC) and impaired participants.

**RESULTS:** There was a significant positive relationship between in-person tPACC and PACC5-RAVLT (ICC=0.88), in-person PACC5-RAVLT and remote tPACC (ICC=0.73), and in-person tPACC and remote tPACC (ICC=0.82). Overall, tPACC showed significant associations with imaging biomarkers.

**DISCUSSION:** There is a good agreement between tPACC and PACC5-RAVLT for NC and impaired individuals. The tPACC is associated with common neuroimaging markers.

**Research in Context:**

**1. Systematic Review:** We reviewed the literature to identify the reliability of remote cognitive testing, concordance of in-person and remote cognitive testing, and remote Preclinical Alzheimer’s Cognitive Composite (PACC) to differentiate cognitive functioning in older adults with or without cognitive impairment. We searched studies to examine relationships between remote PACC score and neuroimaging biomarkers related to early stages of Alzheimer’s Disease. We found no studies simultaneously examining a telehealth PACC (tPACC) score with neuroimaging biomarkers that provide crucial implications for detecting cognitive differences.
**2. Interpretation:** Our findings reveal that a tPACC score can serve as a harmonized composite measure for heterogeneous cognitive data to discriminate those with or without cognitive impairment and inform neuroimaging outcomes.
**3. Future Directions:** Future work is necessary to test tPACC performance in larger longitudinal studies and relationships with other neuroimaging (e.g., PET) or plasma biomarkers in older adults across the cognitive spectrum.

## 1. Introduction

Worldwide, Alzheimer’s disease (AD) and AD related dementias (ADRD) affect over 50 million people, a number likely to triple by 2050^1^. The current clinical trial landscape in ADRD targets individuals with preclinical (i.e., cognitively asymptomatic) disease and mild cognitive impairment (MCI), for whom the hope of averting the pathologic cascade and cognitive decline remains highest. The preclinical Alzheimer’s cognitive composite (PACC) assessment was previously identified to capture early, subtle decline in individuals who are amyloid positive compared to those who are amyloid negative^2, 3^. A popular current version of the PACC, referred to as PACC5^4^, includes five cognitive tests conducted in-person: the Mini-Mental State Examination (MMSE)^5^, Logical Memory Delayed Story Recall (LMDR)^6^, Digit-Symbol Substitution Test (DSST)^6, 7^, recall from the Free and Cued Selective Reminding Test (FCSRT96)^8^, and category fluency^9, 10^. The PACC5 correlates with Amyloid and Fluorodeoxyglucose (FDG) Positron Emission Tomography (PET) imaging^4, 11^. For research and clinical purposes, numerous variations of the PACC have been derived^12–14^. At the Wake Forest Alzheimer’s Disease Research Center (ADRC), for annual Uniform Dataset (UDS)^15, 16^ visits, we assess the PACC5 with minor substitutions (Craft Story Verbatim Delayed Recall^17^ for LMDR, and Rey Auditory Verbal Learning Task^18^ Delayed Recall for FCSRT96), defined as PACC5-RAVLT, only for participants who have visits conducted in-person.

At the outset of the COVID-19 pandemic, in-person cognitive testing was increasingly performed remotely by telephone and video administration^19^, resulting in a mixed (in-clinic and telehealth) cognitive data. Therefore, we developed a harmonized, telehealth-compatible PACC (tPACC) for heterogeneous cognitive data. The tPACC can be derived from in-person UDS v3.0 or UDS v3.0 Tele-Cog^16^ using the following: Montreal Cognitive Assessment (MoCA)^20^, Rey Auditory Verbal Learning Task (RAVLT) Delayed Recall, Craft Story Verbatim Delayed Recall (Craft Story DR) and semantic fluency (i.e., category fluency including animal and vegetables). We calculated a version of tPACC using in-person UDS cognitive data (tPACC-in) in the larger ADRC Clinical Core and remote tPACC (tPACC-rem) using UDS Tele-Cog data that was obtained as part of a smaller validation pilot study for telecognitive assessments. A version of tPACC-in was also generated for pilot study participants. The validation pilot study tPACC scores allowed us to examine mode of administration (in-person vs remote) when forming composite scores.

Though COVID-19 was the initial motivation for the tPACC, remote cognitive assessment can potentially expand research access and reduce participant burden. Given these additional benefits, we anticipate that variation in test modality may become increasingly common independent of COVID-19 trends, amplifying the need for continuity in methods of detecting cognitive change. At the Wake Forest ADRC, remote cognitive assessments are increasingly offered as an alternative to in-person assessments. Consequently, we developed composite measurements similar to previously identified PACC from remotely available assessments to differentiate cognitive status among cognitively normal (NC) and impaired (MCI and dementia) participants.

We evaluated the performance of the tPACC in individuals who are adjudicated as having NC, MCI, or dementia (DEM) to determine whether we can identify subtle cognitive decline. Further, we explored decline in those who have one or two copies of *APOE* ε4 allele, a known risk for AD. We compared the PACC5-RAVLT and tPACC for the agreement in the ADRC Clinical Core cohort and feasibility of tPACC for both in-person and remote cognitive testing in the validation pilot study within the ADRC. Our goals were 1) to evaluate concordance of tPACC scores from in-person and remote testing and 2) to validate them against other known markers of brain and cognitive health. This work is the first to examine the feasibility of a telecognitive-assessment compatible PACC (i.e., tPACC) to be used in longitudinal cohort data collection when administration may vary from in-person to remote (i.e., mixed follow-up data). We will determine whether tPACC can serve as the primary outcome measure for longitudinal studies examining cognitive decline and for predicting common neuroimaging indices.

## 2. Methods

### 2.1. Cohort Characteristics

Our study includes 648 older adults aged 70.0±8.1 years (range: 55-85) who were recruited into the Wake Forest ADRC Clinical Core from the surrounding community between 2016 and 2022. Participants underwent standard evaluation in accordance with the National Alzheimer’s Coordinating Center (NACC)^15^ protocol for neuropsychological data collection using the UDS v3.0^16^ as well as supplementary cognitive tests used at the Wake Forest ADRC (described below). The ADRC Clinical Core cohort is 78% White/Caucasian, 21% Black/African American, and 1% American Indian/Alaska Native or Asian. Participants *APOE* genotyping was done as previously described^21^. Medical history and physical and neurological examinations were completed to rule out major neurologic disorders other than AD. The study was approved by the Wake Forest Institutional Review Board; written protocols and informed consent procedures for all participants and/or their legally authorized representatives were obtained.

We also included 70 older adults (mean age: 71.6±9.1) who were enrolled between late 2020 and 2021 into a validation pilot study within the Wake Forest ADRC investigating the reliability of video/telephone cognitive testing visits in addition to regular visits. Of the 70 participants, 69 were also enrolled in the Clinical Core cohort above. These participants completed additional remote cognitive testing via either telephone or video within six months of their in-person UDS annual assessments. The demographic composition of this sample was comparable to that of the ADRC Clinical Core Cohort described above (80% White and 20% Black/African American). This study was also approved by the Wake Forest Institutional Review Board and informed consent was obtained for all participants, or their representatives, prior to the initiation of study procedures.

### 2.2. Cognitive Groups

All Clinical Core participants undergo adjudication of cognitive status, which includes expert panel consensus review of clinical, neuroimaging, and cognitive data^21^, in accordance with National Institute of Aging-Alzheimer’s Association guidelines for diagnosis of MCI^22^ and DEM. Etiologic diagnosis is made after an adjudicated clinical syndrome^23^. Participants were categorized into NC (adjudicated as NC or NC with subjective complaint) and Impaired (MCI and DEM). Participants adjudicated as “other” and “cognitively impaired not MCI” (CI) were excluded. Additionally, we explored groups by *APOE* ε4 carrier status. Clinical Dementia Rating Sum of Boxes^24, 25^ (CDR-SOB) was examined to confirm cognitive status in the Clinical Core Cohort. Higher CDR-SOB scores indicate impairment in cognitive functioning.

### 2.3. In-person and Remote Cognitive Assessments

Participants’ neuropsychological battery scores for in-person and remote visits were obtained using the UDS v3.0. In-person tests included Montreal Cognitive Assessment (MoCA), Craft Story, Benson Complex Figure, Number Span Test, Verbal Fluency (letters FL), category fluency (Animals and Vegetables), Trail Making Test (TMT), and the Multilingual Naming Test (MINT). In addition, supplementary tests were administered, including Mini-Mental State Exam (MMSE), American National Adult Reading Test (AMNART), Digit Symbol Coding Test (DSC), Free and Cued Selective Reminding Test (FCSRT), Rey Auditory Verbal Learning Test (RAVLT), and Verbal Fluency (letter C). Participants in the pilot telecognitive assessment study underwent a telephone or video administered cognitive testing session in addition to their annual in-person visit. Remote testing included the blind MoCA (omits the Visuospatial/Executive section), Craft Story, RAVLT, Verbal Fluency, Verbal Naming Test (an alternative to MINT), Oral Trail Making Test^26, 27^ (an alternative to written TMT) and Number Span Test.

The composites’ respective component measures are listed in Tables 3 and 4. In this study, we formed a version of PACC5 (PACC5-RAVLT) using RAVLT Delayed Recall (DR) in place of FCSRT96 (for consistency between in-person and remote testing), using in-person cognitive measures, including: RAVLT Delayed Recall (DR), category fluency, Craft Story DR (verbatim), MMSE total, and DSC total. The tPACC included some of the same cognitive measures (RAVLT DR, category fluency and Craft Story DR) for consistency, but replaced MMSE with MoCA, and did not include DSC total as this cannot be administered remotely. We calculated in-person (tPACC-in) and remote tPACC (tPACC-rem) derived from pilot study participant data (n=70). Higher z-scores on the tPACC or PACC5-RAVLT indicate better cognitive performance.

### 2.4. Neuroimaging Data Processing and Analysis

Imaging data acquisition and processing procedures have been described previously^21, 28, 29^. Brain MRI scans were acquired on a 3T Siemens Skyra with a 32-channel head coil (Erlangen, Germany). Anatomical T1-weighted (T1w) and T2-Fluid Attenuated Inversion Recovery (FLAIR), diffusion weighted images for diffusion tensor imaging (DTI) and Neurite orientation dispersion and density imaging (NODDI), and Arterial Spin Labeling (ASL) for whole-brain cerebral blood flow (CBF) were obtained. Structural T1w image processing included normalization and tissue segmentation using SPM12 (www.fil.ion.ucl.ac.uk/spm) CAT12 toolbox. Thickness and volume data on T1w were generated using FreeSurfer v7.2 (http://surfer.nmr.mgh.harvard.edu/). We calculated cortical thickness in AD-vulnerable regions from a temporal lobe meta region of interest (Temporal Meta ROI) by averaging cortical thickness of bilateral entorhinal, inferior/middle temporal, and fusiform regions, as well as hippocampal volume (HCV) expressed as % of Intracranial Volume (ICV). White matter hyperintensities (WMH) were segmented by Lesion Segmentation Tool (LST) v2.0.15. (https://www.applied-statistics.de/lst.html), log-transformed and scaled to ICV for analysis (Log-WMH). The Johns Hopkins University (JHU) DTI atlas^30^ was overlaid on template-space fractional anisotropy (FA) and free water (FW) maps to extract mean signal across all supratentorial white matter (WM) tracts. Automated Anatomical Labeling (AAL)^31^ gray matter (GM) ROIs were overlaid on template-space GM CBF images to calculate mean hippocampal, frontal lobe, and a temporal meta-ROI (parahippocampus, fusiform, middle, and inferior temporal cortex) GM CBF, and a set of all supratentorial JHU WM tracts were overlaid on template-space WM CBF images to calculate mean global WM CBF.

### 2.5. Statistical Analyses

All statistical analyses were performed in SAS v9.4. We explored group differences for baseline demographics, cognitive measures and neuroimaging based on adjudicated cognitive status in the ADRC Clinical Core cohort and the Validation Pilot Study cohort. Additionally, we examined PACC5-RAVLT and tPACC by *APOE* ε4 carrier status. We conducted Intraclass Correlation Coefficient Analysis (ICC) to describe agreement between PACC5-RAVLT and tPACC in all baseline in-person visits. We then generated Bland-Altman Plots to visualize agreement between these two measures. Spearman correlation was used for MMSE and MoCA and for RAVLT and FCSRT96 to assess their relationships. We repeated ICC and Bland-Altman analyses in pilot study individuals who completed tPACC-in and tPACC-rem within 6 months (median: 151 days; IQR: 110, 288) to evaluate the reliability between in-person and remote administration. General linear models were used to examine baseline neuroimaging markers as dependent variables and tPACC scores as independent variables, adjusting for age, sex, education years, race (self-report; social construct variable) and *APOE* ε4, in ADRC Clinical Core participants. Statistical significance for linear models was *P* < 0.05.

## 3. Results

We included a total of 649 older participants from two cohorts, of whom 334 (51%) were NC and 315 (49%) were cognitively impaired (See Tables 1 and 2 for a breakdown by cohort). All impaired groups tended to be older and have lower education years and higher CDR-SOB scores than NC. There were no sex differences between groups. A majority were White participants. Of all, 32% had one or two copies of *APOE*4.

**Table 1:**
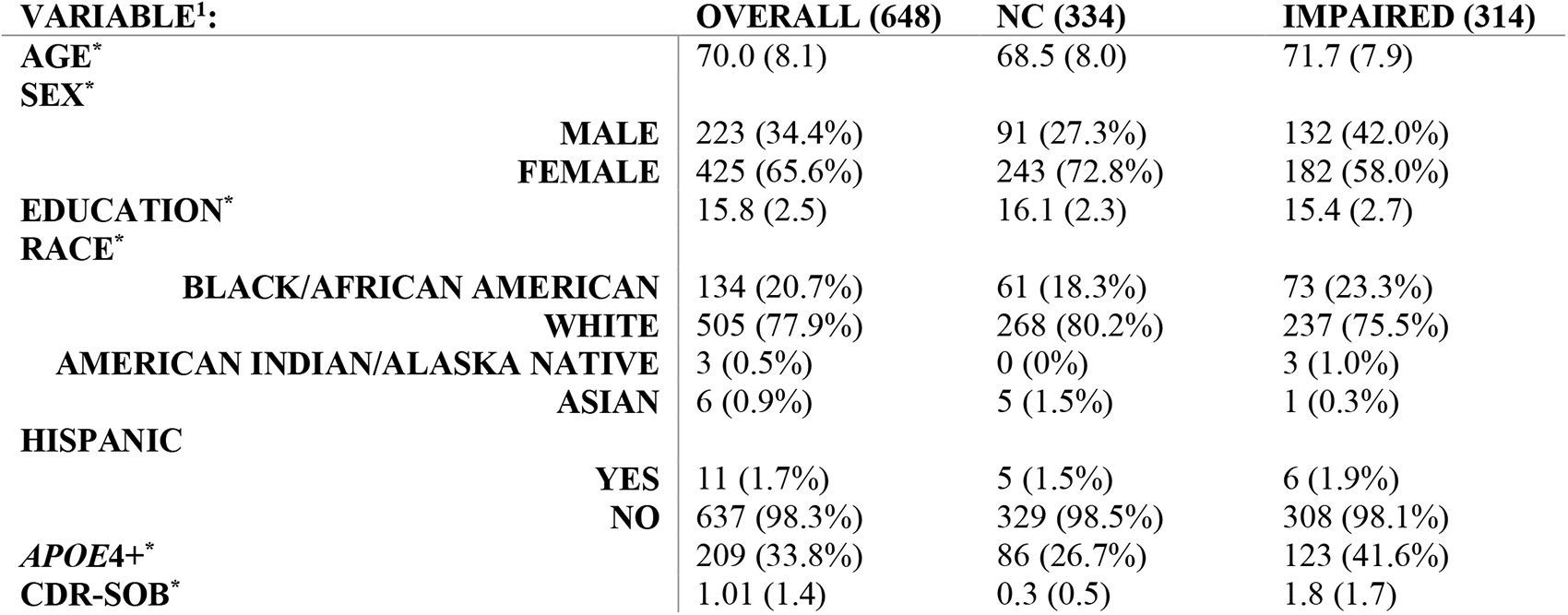
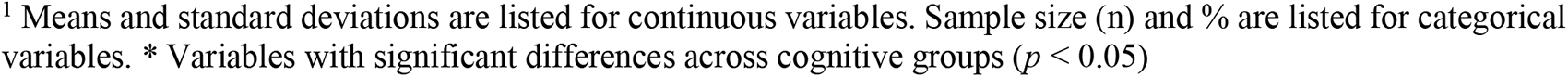
Demographic Characteristics by cognitive groups in the ADRC Clinical Core Cohort.

| **VARIABLE<sup>1</sup>:** |  | **OVERALL (648)** | **NC (334)** | **IMPAIRED (314)** |
| --- | --- | --- | --- | --- |
| **AGE\*** |  | 70.0 (8.1) | 68.5 (8.0) | 71.7 (7.9) |
| **SEX\*** |  |  |  |  |
|  | **MALE** | 223 (34.4%) | 91 (27.3%) | 132 (42.0%) |
|  | **FEMALE** | 425 (65.6%) | 243 (72.8%) | 182 (58.0%) |
| **EDUCATION\*** |  | 15.8 (2.5) | 16.1 (2.3) | 15.4 (2.7) |
| **RACE\*** |  |  |  |  |
|  | **BLACK/AFRICAN AMERICAN** | 134 (20.7%) | 61 (18.3%) | 73 (23.3%) |
|  | **WHITE** | 505 (77.9%) | 268 (80.2%) | 237 (75.5%) |
|  | **AMERICAN INDIAN/ALASKA NATIVE** | 3 (0.5%) | 0 (0%) | 3 (1.0%) |
|  | **ASIAN** | 6 (0.9%) | 5 (1.5%) | 1 (0.3%) |
| **HISPANIC** |  |  |  |  |
|  | **YES** | 11 (1.7%) | 5 (1.5%) | 6 (1.9%) |
|  | **NO** | 637 (98.3%) | 329 (98.5%) | 308 (98.1%) |
| **APOE4+\*** |  | 209 (33.8%) | 86 (26.7%) | 123 (41.6%) |
| **CDR-SOB\*** |  | 1.01 (1.4) | 0.3 (0.5) | 1.8 (1.7) |

**Table 2:**
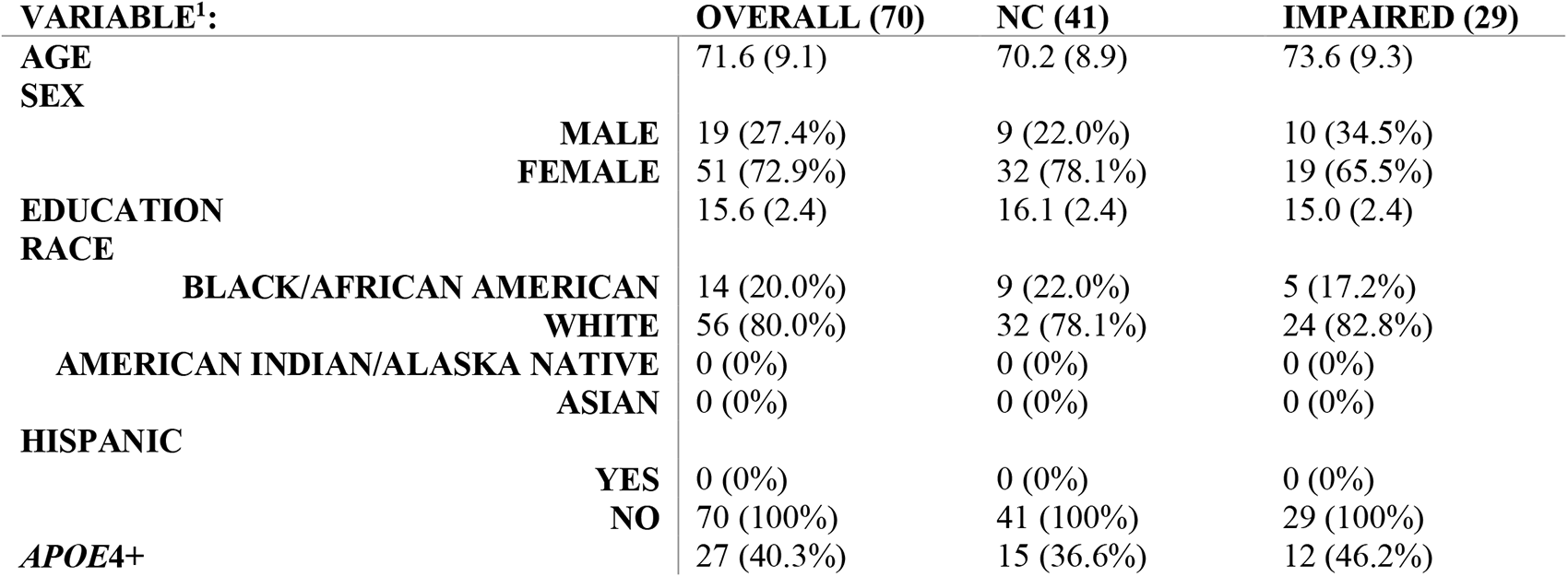
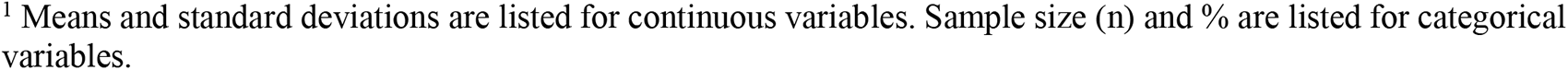
Demographic Characteristics in the Validation Pilot Study Cohort.

| **VARIABLE<sup>1</sup>:** |  | **OVERALL (70)** | **NC (41)** | **IMPAIRED (29)** |
| --- | --- | --- | --- | --- |
| **AGE** |  | 71.6 (9.1) | 70.2 (8.9) | 73.6 (9.3) |
| **SEX** |  |  |  |  |
|  | **MALE** | 19 (27.4%) | 9 (22.0%) | 10 (34.5%) |
|  | **FEMALE** | 51 (72.9%) | 32 (78.1%) | 19 (65.5%) |
| **EDUCATION** |  | 15.6 (2.4) | 16.1 (2.4) | 15.0 (2.4) |
| **RACE** |  |  |  |  |
|  | **BLACK/AFRICAN AMERICAN** | 14 (20.0%) | 9 (22.0%) | 5 (17.2%) |
|  | **WHITE** | 56 (80.0%) | 32 (78.1%) | 24 (82.8%) |
|  | **AMERICAN INDIAN/ALASKA NATIVE** | 0 (0%) | 0 (0%) | 0 (0%) |
|  | **ASIAN** | 0 (0%) | 0 (0%) | 0 (0%) |
| **HISPANIC** |  |  |  |  |
|  | **YES** | 0 (0%) | 0 (0%) | 0 (0%) |
|  | **NO** | 70 (100%) | 41 (100%) | 29 (100%) |
| **APOE4+** |  | 27 (40.3%) | 15 (36.6%) | 12 (46.2%) |

<sup>1</sup> Means and standard deviations are listed for continuous variables. Sample size (n) and % are listed for categorical variables. \* Variables with significant differences across cognitive groups ( $p < 0.05$ )

Spearman correlation coefficients for MoCA with MMSE and RAVLT with FCSRT96 were .63 and .69, respectively. Baseline cognitive measures used in PACC5-RAVLT and tPACC scores were lower/worse in impaired groups in both cohorts (Tables 3 and 4). In person PACC5-RAVLT and tPACC scores were lower in impaired and *APOE*4+ (Tables 5 and 6). Bland-Altman plots revealed a good level of agreement between tPACC and PACC5-RAVLT (Figures 1-4). On average, 98% of all values lie within limit of agreement (±2SD and ±3SD), indicating good reliability (overall ICCs=.88, .90, .73, and .82, respectively) between tPACC and PACC5-RAVLT in each group. Furthermore, an overestimation of tPACC was observed in the impaired group (Figure 1).

**Table 3:**
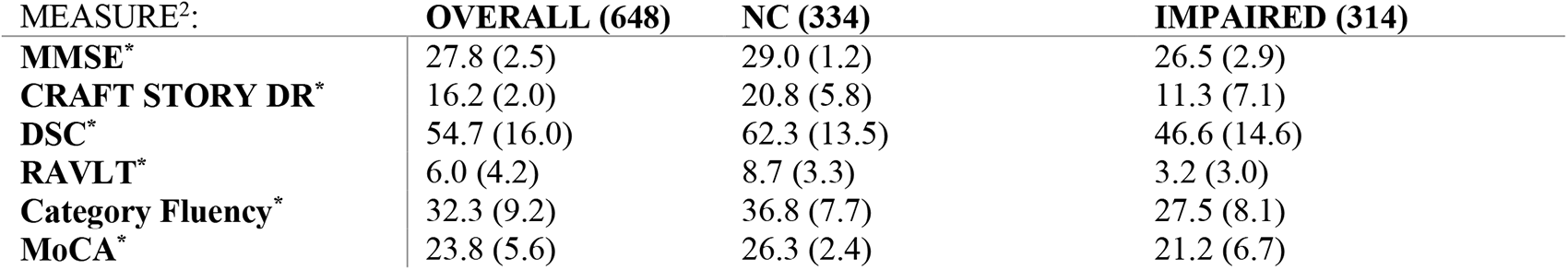
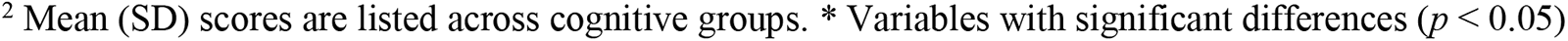
Baseline scores of cognitive measures used in PACC5-RAVLT and tPACC in the ADRC Clinical Core Cohort.

| MEASURE<sup>2</sup>: | OVERALL (648) | NC (334) | IMPAIRED (314) |
| --- | --- | --- | --- |
| MMSE* | 27.8 (2.5) | 29.0 (1.2) | 26.5 (2.9) |
| CRAFT STORY DR* | 16.2 (2.0) | 20.8 (5.8) | 11.3 (7.1) |
| DSC* | 54.7 (16.0) | 62.3 (13.5) | 46.6 (14.6) |
| RAVLT* | 6.0 (4.2) | 8.7 (3.3) | 3.2 (3.0) |
| Category Fluency* | 32.3 (9.2) | 36.8 (7.7) | 27.5 (8.1) |
| MoCA* | 23.8 (5.6) | 26.3 (2.4) | 21.2 (6.7) |

**Table 4:**
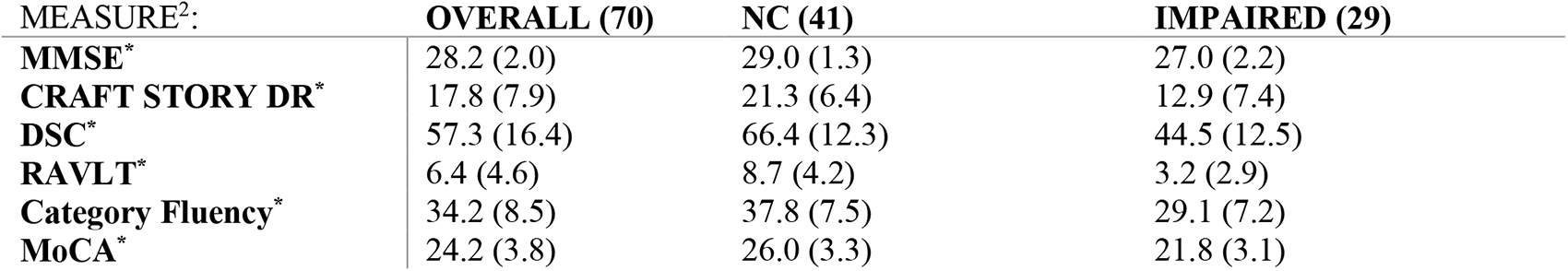
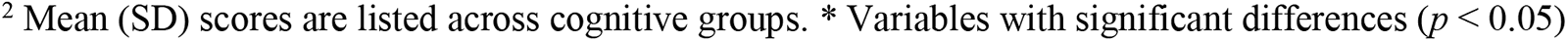
Baseline scores of cognitive measures used in PACC5-RAVLT and tPACC in the Validation Pilot Study.

| MEASURE<sup>2</sup>: | OVERALL (70) | NC (41) | IMPAIRED (29) |
| --- | --- | --- | --- |
| MMSE* | 28.2 (2.0) | 29.0 (1.3) | 27.0 (2.2) |
| CRAFT STORY DR* | 17.8 (7.9) | 21.3 (6.4) | 12.9 (7.4) |
| DSC* | 57.3 (16.4) | 66.4 (12.3) | 44.5 (12.5) |
| RAVLT* | 6.4 (4.6) | 8.7 (4.2) | 3.2 (2.9) |
| Category Fluency* | 34.2 (8.5) | 37.8 (7.5) | 29.1 (7.2) |
| MoCA* | 24.2 (3.8) | 26.0 (3.3) | 21.8 (3.1) |

**Figure 1:**
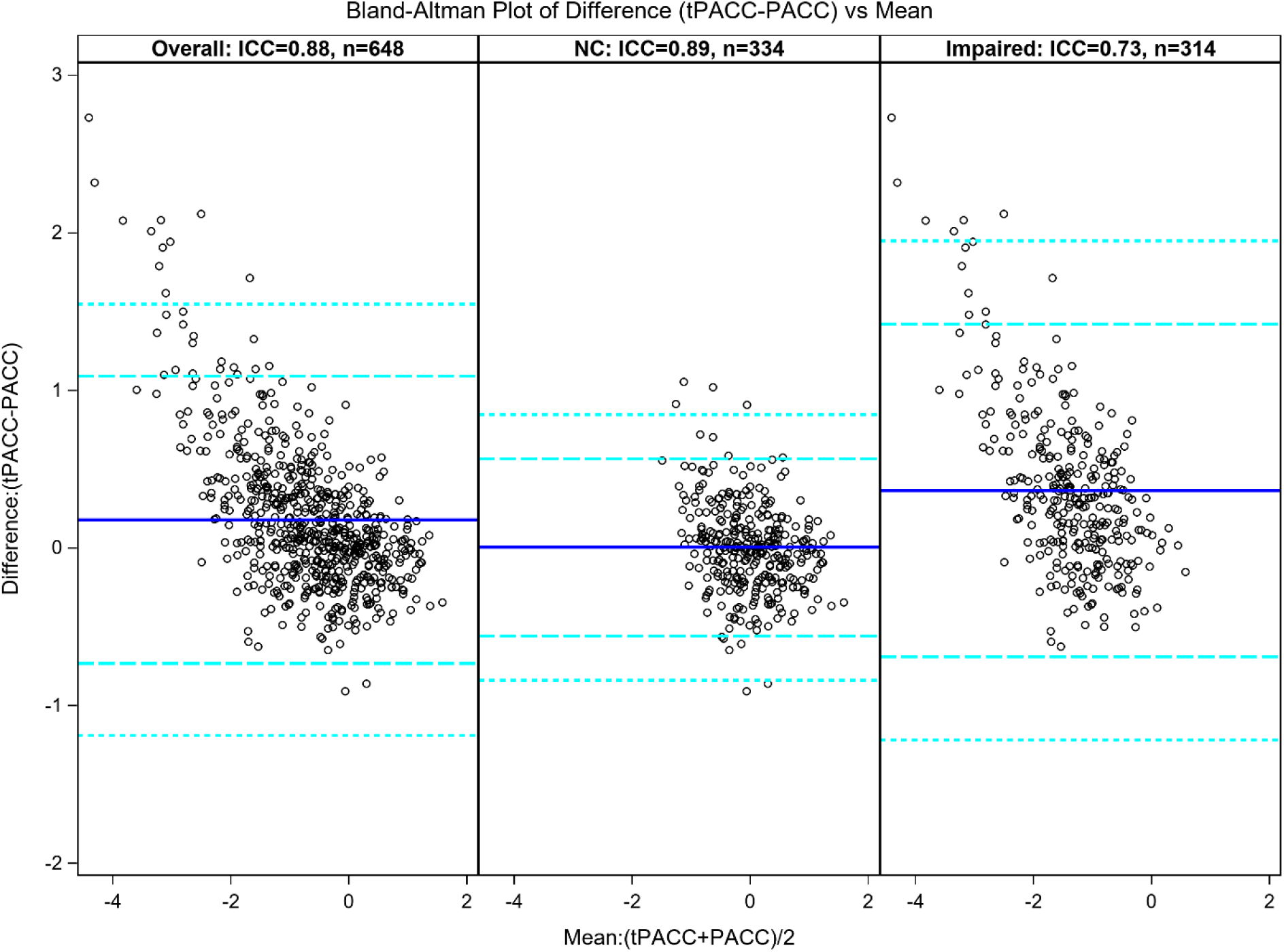
tPACC-in vs PACC5-RAVLT in ADRC Clinical Core Cohort.

**Figure 2:**
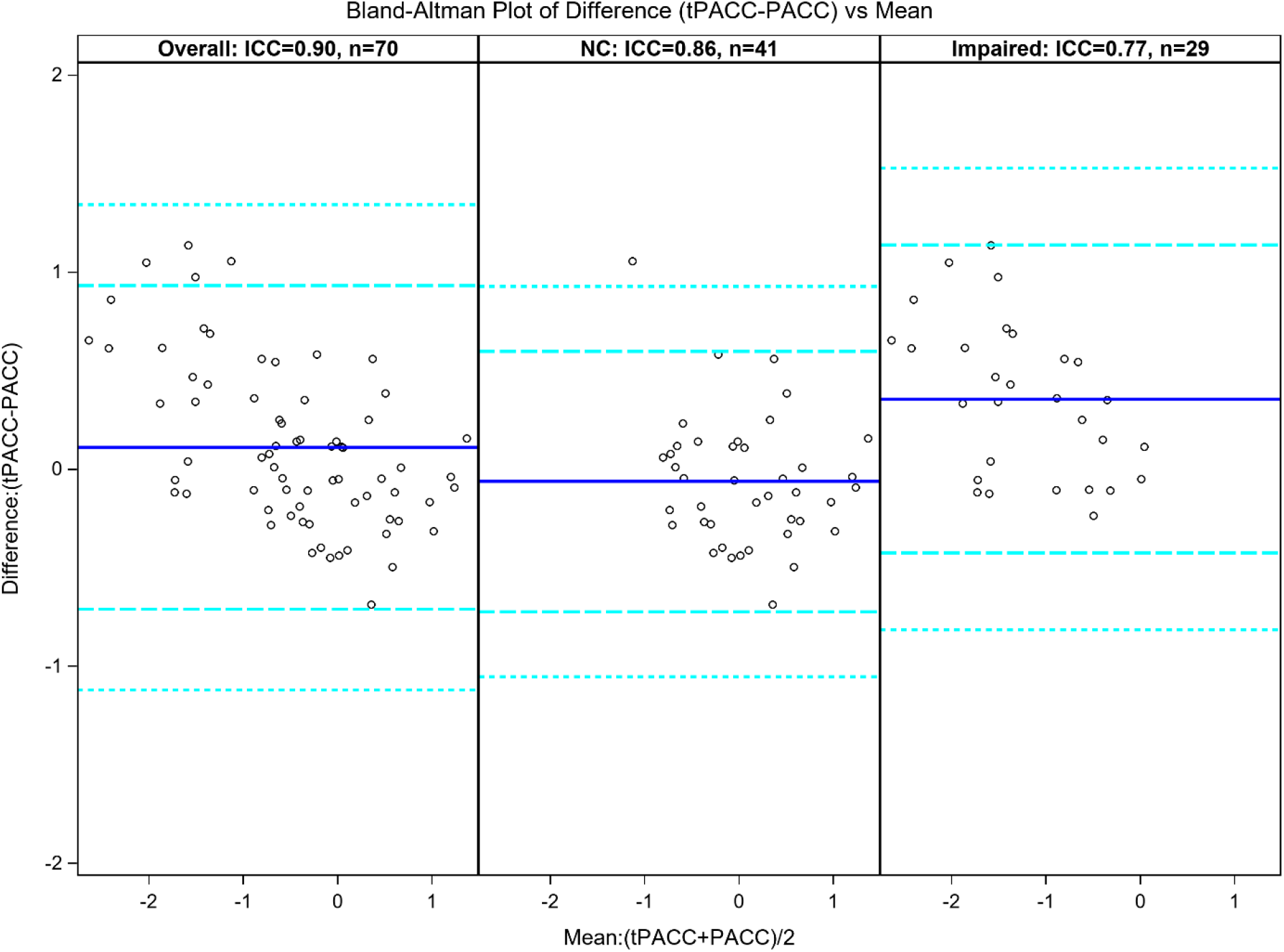
tPACC-in vs PACC5-RAVLT in Validation Pilot Study Cohort.

**Figure 3:**
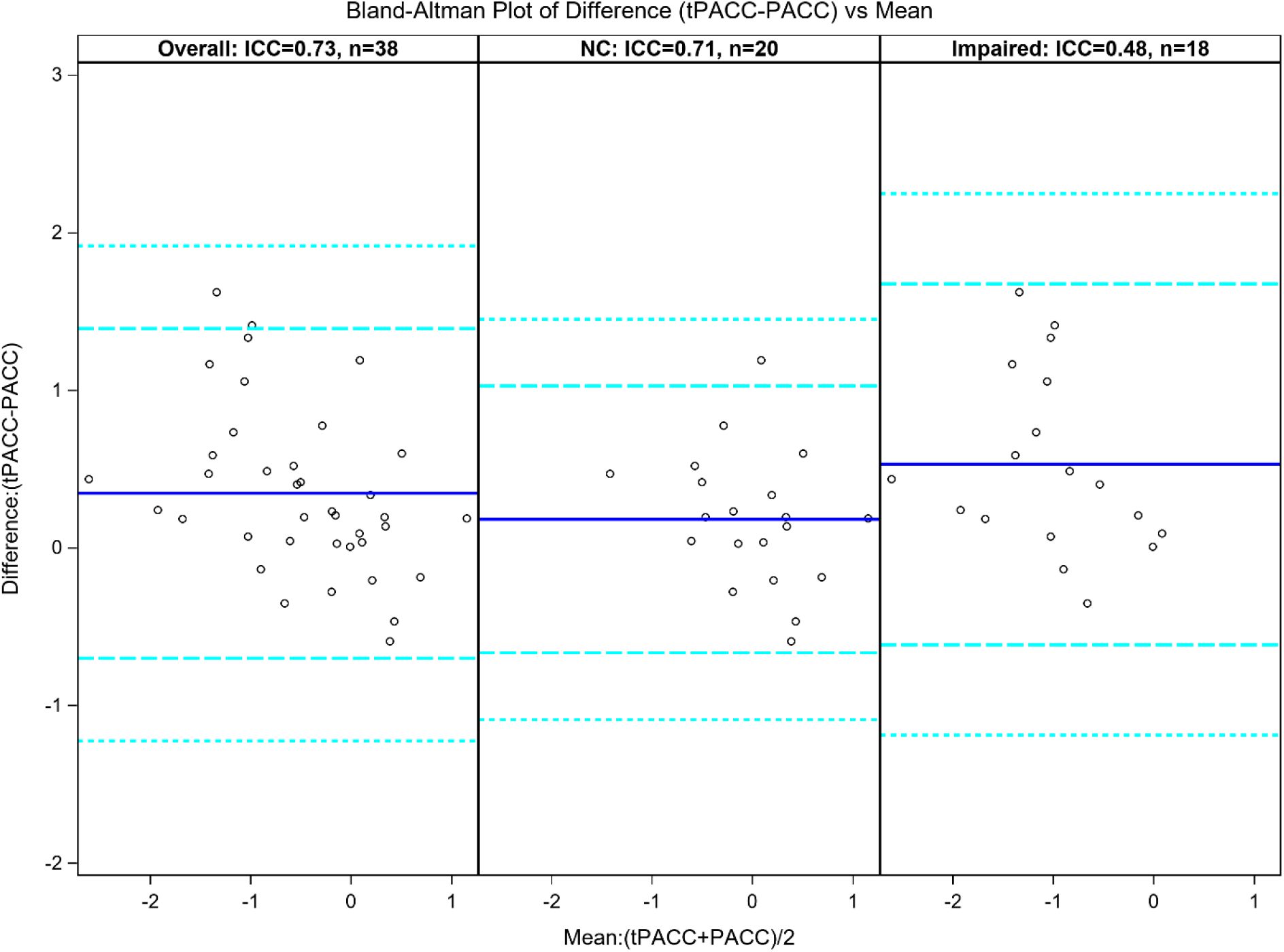
tPACC-rem vs PACC5-RAVLT in Validation Pilot Study Cohort.

**Figure 4:**
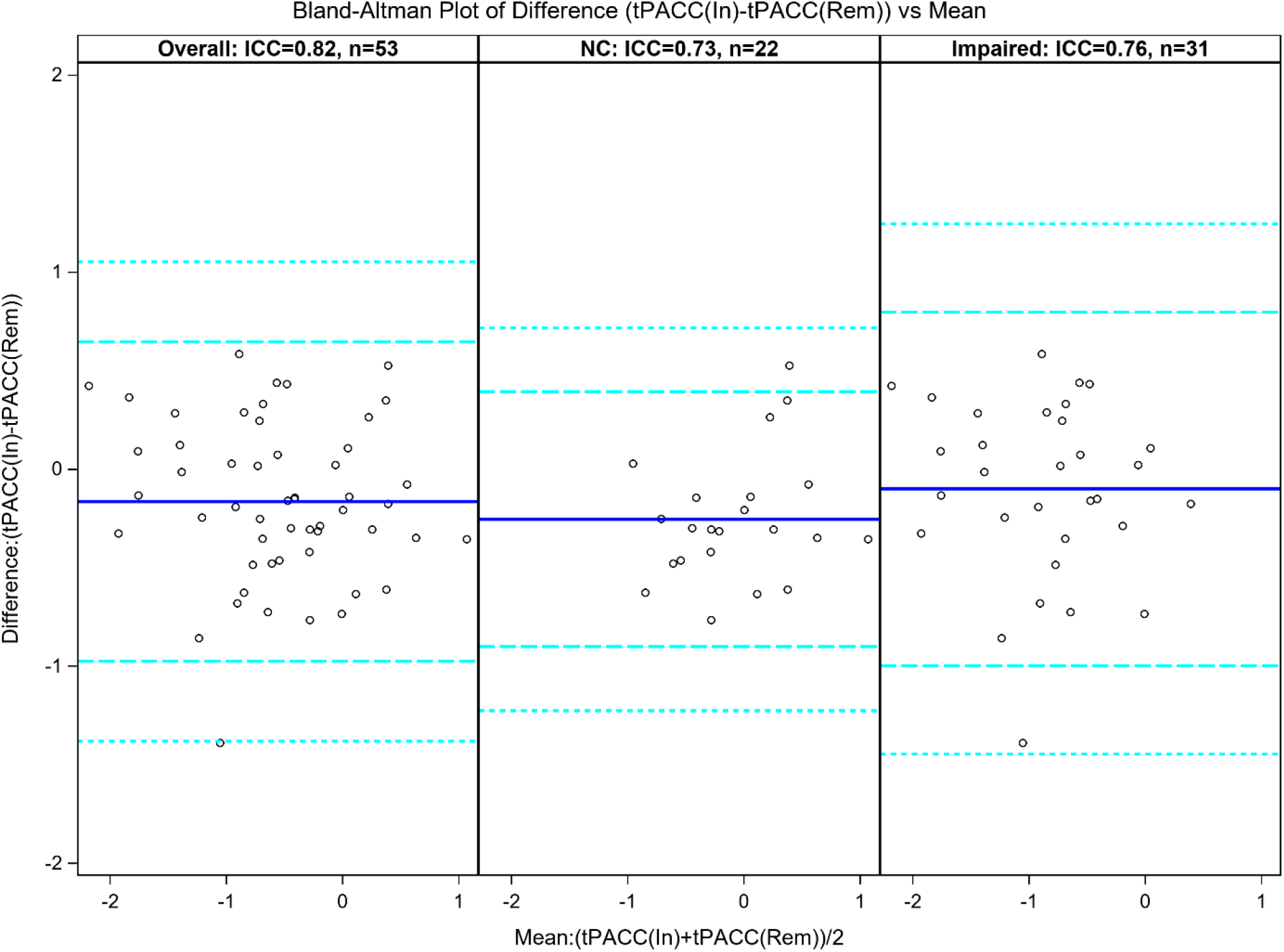
tPACC-rem vs tPACC-in in Validation Pilot Study Cohort.

General linear models adjusting for age, sex, education, race and *APOE*4 indicated that tPACC showed significant associations with whole brain volume (adjusted for ICV), Temporal Meta ROI thickness, HCV, Log-WMH, ASL CBF (in hippocampi, temporal Meta ROI, and frontal GM) and NODDI WM FW in the overall ADRC Clinical Core cohort (Table 7). Lower tPACC scores were associated with lower brain volume, cortical thickness, HCV and CBF, and higher WMH and FW in WM.

**Table 5:**
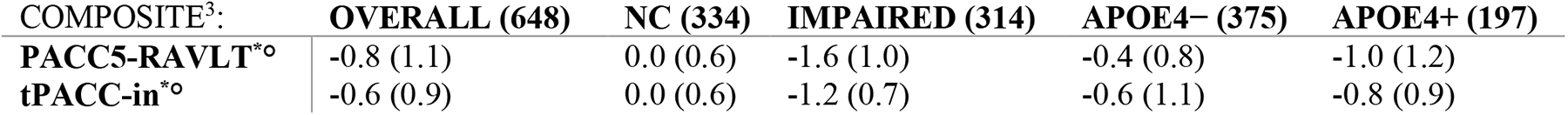
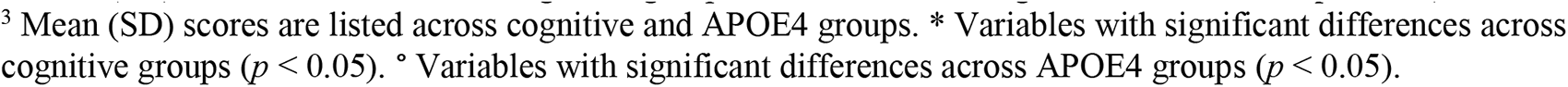
ADRC Clinical Core Cohort PACC5-RAVLT and tPACC-in by cognitive groups and APOE4 status.

| COMPOSITE<sup>3</sup>: | OVERALL (648) | NC (334) | IMPAIRED (314) | APOE4- (375) | APOE4+ (197) |
| --- | --- | --- | --- | --- | --- |
| PACC5-RAVLT*<sup>o</sup> | -0.8 (1.1) | 0.0 (0.6) | -1.6 (1.0) | -0.4 (0.8) | -1.0 (1.2) |
| tPACC-in*<sup>o</sup> | -0.6 (0.9) | 0.0 (0.6) | -1.2 (0.7) | -0.6 (1.1) | -0.8 (0.9) |

**Table 6:**
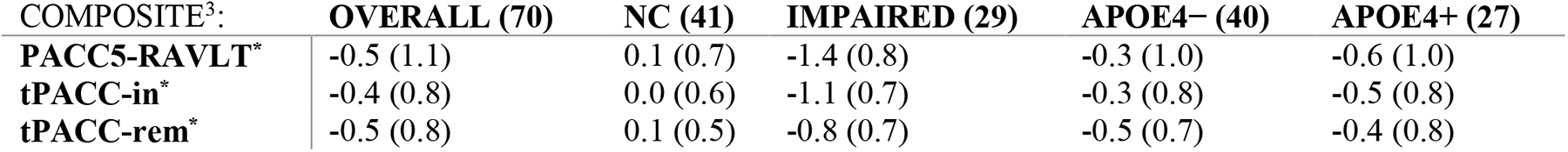
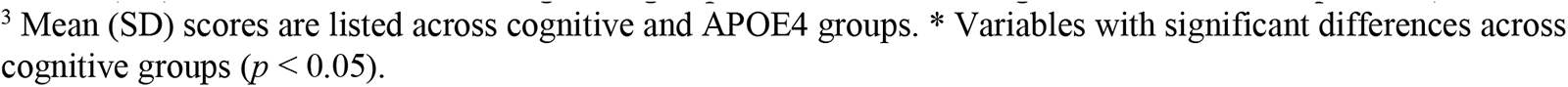
Validation Pilot Study Cohort PACC5-RAVLT and tPACC by cognitive groups and APOE4 status.

| COMPOSITE<sup>3</sup>: | OVERALL (70) | NC (41) | IMPAIRED (29) | APOE4- (40) | APOE4+ (27) |
| --- | --- | --- | --- | --- | --- |
| PACC5-RAVLT* | -0.5 (1.1) | 0.1 (0.7) | -1.4 (0.8) | -0.3 (1.0) | -0.6 (1.0) |
| tPACC-in* | -0.4 (0.8) | 0.0 (0.6) | -1.1 (0.7) | -0.3 (0.8) | -0.5 (0.8) |
| tPACC-rem* | -0.5 (0.8) | 0.1 (0.5) | -0.8 (0.7) | -0.5 (0.7) | -0.4 (0.8) |

<sup>2</sup> Mean (SD) scores are listed across cognitive groups. \* Variables with significant differences ( $p < 0.05$ )
<sup>3</sup> Mean (SD) scores are listed across cognitive and APOE4 groups. \* Variables with significant differences across cognitive groups ( $p < 0.05$ ). <sup>o</sup> Variables with significant differences across APOE4 groups ( $p < 0.05$ ).

**Table 7:** Neuroimaging markers (outcome) and tPACC.

| OUTCOME | Beta (95% Confidence Interval) | *p*-value |
| --- | --- | --- |
| Temporal Meta ROI Thickness, mm | 0.07 (0.06, 0.09) | <0.0001 |
| Whole Brain Volume, L | 0.01 (0.01, 0.02) | <0.0001 |
| HCV, % of ICV | 0.03 (0.02, 0.04) | <0.0001 |
| Log-WMH | -0.20 (-0.30, -0.10) | 0.0002 |
| ASL CBF Hippocampi | 1.28 (0.24, 2.32) | 0.0161 |
| ASL CBF Temporal Meta ROI | 1.43 (0.61, 2.26) | 0.0007 |
| ASL CBF Frontal Lobe | 1.04 (0.24, 1.84) | 0.0113 |
| ASL CBF WM | 0.10 (-0.31, 0.51) | 0.6361 |
| DTI_FA WM | 0.0002 (-0.0024, 0.0028) | 0.8880 |
| NODDI WM FW | -0.0048 (-0.0075, -0.0020) | 0.0007 |

**Table 7.1:** Neuroimaging markers and tPACC (outcome)

| VARIABLE | Beta (95% Confidence Interval) | *p*-value |
| --- | --- | --- |
| Temporal Meta ROI Thickness | 1.87 (1.47, 2.27) | <0.0001 |
| Whole Brain Volume | 5.81 (4.04, 7.59) | <0.0001 |
| HCV | 4.15 (3.24, 5.06) | <0.0001 |
| Log-WMH | -0.13 (-0.20, -0.06) | 0.0002 |
| ASL CBF Hippocampi | 0.0087 (0.0016, 0.0157) | 0.0161 |
| ASL CBF Temporal Meta ROI | 0.02 (0.01, 0.02) | 0.0007 |
| ASL CBF Frontal Lobe | 0.0118 (0.0027, 0.0209) | 0.0113 |
| ASL CBF WM | 0.0043 (-0.014, 0.0223) | 0.6361 |
| DTI_FA WM | 0.21 (-2.74, 3.17) | 0.8880 |
| NODDI WM FW | -5.06 (-7.98, -2.14) | 0.0007 |

## Discussion

To examine whether a telehealth derived PACC (tPACC) can be used for both in-person and remote visits to reflect cognitive function, we performed a reliability study in 649 participants from two Wake Forest ADRC cohorts with in-person and remote cognitive data available. As expected, we found that tPACC performs similarly to in-person PACC5-RAVLT with ICCs ranging from .73 to .90; tPACC thus may serve as a reliable measure in place of the PACC5-RAVLT for heterogeneous test administration settings. Our findings revealed that tPACC differentiates cognitive performance levels across NC and impaired individuals. Furthermore, neuroimaging biomarkers (Temporal Meta ROI thickness, HCV, WMH, CBF, and WM FW) showed significant associations with tPACC among Clinical Core participants. Taken together, these findings suggest that tPACC, when given in-person or remotely, can be used for mixed study visits (an important finding given the increased interest in remote assessments), and can also demonstrate associations with established neuroimaging biomarkers of AD-related cognitive impairment.

The PACC is designed to measure objective cognitive performance in AD-related domains, including memory, executive functioning, attention, and language, and to potentially serve as a primary endpoint for preclinical AD trials. It is typically composed of a battery of neuropsychological tests administered at in-person research study visits. Since the 2020 COVID-19 pandemic affected the ability of research centers to conduct in-person clinical research study visits, remote testing has increasingly become more accessible, affordable, convenient and less burdensome to older participants^32, 33^. Research has shown that older individuals with better PACC scores tend to have a lower risk for AD over time^12, 34, 35^. It is crucial to monitor cognitive performance regardless of mode of administration using a harmonized PACC that is similarly sensitive to detect subtle cognitive changes, in order to increase accessibility to and validity of research studies investigating late-life cognitive health.

Previous studies have shown feasibility and validity of remote cognitive testing including both telephone and video-administered assessments for older adults^36, 37^. Most research participants are comfortable using technology (device, internet etc.) for assessments. The results from remote cognitive testing are comparable to in-person testing. In general, there is high concordance between in-person and remote assessments^38, 39^. However, studies also noted slight differences for certain cognitive domain tests (e.g., complex visuospatial tasks) when administered by video or telephone. Overall research shows that remote testing is a feasible and reliable alternative to in-person testing and provides acceptable results for researchers and convenience and accessibility to participants.

Evaluating the reliability of tPACC derived from in-person and remote testing is necessary for longitudinal studies that examine neuropsychological testing administered in heterogeneous settings (e.g., in-clinic vs remotely). Importantly, knowledge is limited regarding how tPACC can be derived from remotely administered cognitive assessments, and how strongly remote PACC measures like tPACC are associated with common AD-related neuroimaging biomarkers. A 2021 study by Seghezzo et al.^40^ showed the feasibility of conducting remote PACC via videoconferencing and the use of videoconferencing when in-person assessments cannot be done. Similarly, the current study provides evidence for feasibility of remote testing and validates how similar the PACC scores via in-person vs remote.

Our findings extend these data for tPACC by demonstrating significant associations with known neuroimaging biomarkers. While previously identified PACC scores were significantly associated with amyloid-related decline assessed by Amyloid PET^3^, we extent this relationship of PACC and neuroimaging by using a harmonized measure and MRI markers implicated in AD. The use of tPACC and common AD MRI measures has crucial implications in early detection and diagnosis, prediction of disease progression, evaluation of treatment effectiveness, identifying at-risk individuals, and AD research. These measures contribute to our understanding of the preclinical disease and have the potential to improve clinical trial outcomes.

Major strengths of the current study include our ability to examine the reliability of in-person and remote administration modes in a relatively large clinical cohort, and the use of neuroimaging to further understand relationships between the tPACC and multiple MRI indices. Some limitations of our study are worth noting. The sample size from the validation pilot study was relatively small compared to larger studies investigating remote neuropsychological assessments. All observations made here are cross-sectional and need to be further replicated in the general population in longitudinal studies. Although we have limited inclusion of underrepresented racial groups in the current samples, the Wake Forest ADRC Clinical Core cohort is ongoing and actively recruiting participants from underrepresented groups. There is also a potential for variability in the participants’ technological resources and familiarity with the technology; however, accumulating evidence shows that older adults are increasingly interested in use of technology, technological devices and internet for research studies. Lastly, there is a potential for learning and practice effects (especially common for memory tests) in the validation pilot study cohort, and our study did not correct for these effects (no randomization for the order of administration modality). Lastly, we do not address the reliability of longitudinal changes and how well tPACC can detect cognitive decline over time. This will be addressed in our future work.

In this study, we defined a telehealth compatible PACC measures (tPACC) and examined its concordance with PACC5-RAVLT. We then focused on the relationships between tPACC and specific AD-related neuroimaging measures, including Temporal Meta ROI thickness, HCV, WMH, CBF, and WM FW. Future directions include determining whether we can identify subtle cognitive decline in those who are amyloid positive (by PET, CSF, or plasma when available) and have subjective cognitive decline (defined by the Cognitive Change Index: CCI^41^), or mild behavioral impairment (defined by the Neuropsychiatric Inventory Questionnaire: NPIQ^42^, or Mild Behavioral Impairment Checklist: MBI-C^43^), or a family history of AD in a first-degree relative. To further understand whether tPACC can detect AD-related cognitive and behavioral decline, we will examine the performance of tPACC in a longitudinal study.

## Data Availability

Final data derived from this study will be shared via publication of the results in a peer-reviewed
Journal. In addition, we will provide guidance, code, protocols, and assistance to any researcher or other interested individuals who contact us regarding our published work.

https://www.wakeshare.org/

## REFERENCES

1. Collaborators GBDDF. Estimation of the global prevalence of dementia in 2019 and forecasted prevalence in 2050: an analysis for the Global Burden of Disease Study 2019. Lancet Public Health. Feb 2022;7(2):e105–e125. doi:10.1016/S2468-2667(21)00249-8

2. Donohue MC, Sperling RA, Salmon DP, et al. The Preclinical Alzheimer Cognitive Composite. JAMA Neurology. 2014;71(8):961. doi:10.1001/jamaneurol.2014.803

3. Donohue MC, Sperling RA, Salmon DP, et al. The preclinical Alzheimer cognitive composite: measuring amyloid-related decline. JAMA Neurol. Aug 2014;71(8):961–70. doi:10.1001/jamaneurol.2014.803

4. Papp KV, Rentz DM, Orlovsky I, Sperling RA, Mormino EC. Optimizing the preclinical Alzheimer’s cognitive composite with semantic processing: The PACC5. Alzheimers Dement (N Y). Nov 2017;3(4):668–677. doi:10.1016/j.trci.2017.10.004

5. Folstein MF, Folstein SE, McHugh PR. “Mini-mental state”: A practical method for grading the cognitive state of patients for the clinician. Journal of Psychiatric Research. 1975/11/01/ 1975;12(3):189–198. doi:https://doi.org/10.1016/0022-3956(75)90026-6

6. Wechsler D. Wechsler adult intelligence scale-revised (WAIS-R). Psychological Corporation; 1981.

7. Jaeger J. Digit Symbol Substitution Test: The Case for Sensitivity Over Specificity in Neuropsychological Testing. J Clin Psychopharmacol. Oct 2018;38(5):513–519. doi:10.1097/jcp.0000000000000941

8. Grober E, Ocepek-Welikson K, Teresi JA. The free and cued selective reminding test: evidence of psychometric adequacy. Psychology Science Quarterly. 2009;51(3):266–282.

9. Morris JC, Heyman A, Mohs RC, et al. The Consortium to Establish a Registry for Alzheimer’s Disease (CERAD). Part I. Clinical and neuropsychological assessment of Alzheimer’s disease. Neurology. Sep 1989;39(9):1159–65. doi:10.1212/wnl.39.9.1159

10. 10. Weintraub S, Salmon D, Mercaldo N, et al. The Alzheimer’s Disease Centers’ Uniform Data Set (UDS): the neuropsychologic test battery. Alzheimer Dis Assoc Disord. Apr-Jun 2009;23(2):91–101. doi:10.1097/WAD.0b013e318191c7dd

11. Mayblyum DV, Becker JA, Jacobs HIL, et al. Comparing PET and MRI Biomarkers Predicting Cognitive Decline in Preclinical Alzheimer Disease. Neurology. May 5 2021;96(24):e2933–43. doi:10.1212/WNL.0000000000012108

12. Mormino EC, Papp KV, Rentz DM, et al. Early and late change on the preclinical Alzheimer’s cognitive composite in clinically normal older individuals with elevated amyloid β. Alzheimer’s & Dementia. 2017;13(9):1004–1012. doi:10.1016/j.jalz.2017.01.018

13. Hampton OL, Mukherjee S, Properzi MJ, et al. Harmonizing the preclinical Alzheimer cognitive composite for multi-cohort studies. Alzheimer’s & Dementia. 2020;16(S9)doi:10.1002/alz.047423

14. Stricker NH, Twohy EL, Albertson SM, et al. Mayo-PACC: A parsimonious preclinical Alzheimer’s disease cognitive composite comprised of public-domain measures to facilitate clinical translation. Alzheimer’s & Dementia. 2022;doi:10.1002/alz.12895

15. 15. Beekly DL, Ramos EM, Lee WW, et al. The National Alzheimer’s Coordinating Center (NACC) database: the Uniform Data Set. Alzheimer Dis Assoc Disord. Jul-Sep 2007;21(3):249–58. doi:10.1097/WAD.0b013e318142774e

16. 16. Weintraub S, Besser L, Dodge HH, et al. Version 3 of the Alzheimer Disease Centers’ Neuropsychological Test Battery in the Uniform Data Set (UDS). Alzheimer Dis Assoc Disord. Jan-Mar 2018;32(1):10–17. doi:10.1097/WAD.0000000000000223

17. Kaur A, Edland SD, Peavy GM. The MoCA-Memory Index Score. Alzheimer Disease & Associated Disorders. 2018;32(2):120–124. doi:10.1097/wad.0000000000000240

18. Bean J. Rey Auditory Verbal Learning Test, Rey AVLT. In: Kreutzer JS, DeLuca J, Caplan B, eds. Encyclopedia of Clinical Neuropsychology. Springer New York; 2011:2174–2175.

19. Nunnerley M, Mattek N, Kaye J, Beattie Z. Preferences of NIA Alzheimer’s Disease Research Center participants regarding remote assessment during the COVID-19 pandemic. *Alzheimer’s & Dementia: Diagnosis*, Assessment & Disease Monitoring. 2022;14(1)doi:10.1002/dad2.12373

20. Nasreddine ZS, Phillips NA, Bã©Dirian VR, et al. The Montreal Cognitive Assessment, MoCA: A Brief Screening Tool For Mild Cognitive Impairment. Journal of the American Geriatrics Society. 2005;53(4):695–699. doi:10.1111/j.1532-5415.2005.53221.x

21. Hughes TM, Lockhart SN, Suerken CK, et al. Hypertensive Aspects of Cardiometabolic Disorders Are Associated with Lower Brain Microstructure, Perfusion, and Cognition. J Alzheimers Dis. Oct 28 2022;doi:10.3233/JAD-220646

22. Albert MS, Dekosky ST, Dickson D, et al. The diagnosis of mild cognitive impairment due to Alzheimer’s disease: Recommendations from the National Institute on Aging-Alzheimer’s Association workgroups on diagnostic guidelines for Alzheimer’s disease. Alzheimer’s & Dementia. 2011;7(3):270–279. doi:10.1016/j.jalz.2011.03.008

23. McKhann GM, Knopman DS, Chertkow H, et al. The diagnosis of dementia due to Alzheimer’s disease: Recommendations from the National Institute on Aging-Alzheimer’s Association workgroups on diagnostic guidelines for Alzheimer’s disease. Alzheimer’s & Dementia. 2011;7(3):263–269. doi:10.1016/j.jalz.2011.03.005

24. Morris JC. The Clinical Dementia Rating (CDR): current version and scoring rules. Neurology. Nov 1993;43(11):2412–4. doi:10.1212/wnl.43.11.2412-a

25. Cedarbaum JM, Jaros M, Hernandez C, et al. Rationale for use of the Clinical Dementia Rating Sum of Boxes as a primary outcome measure for Alzheimer’s disease clinical trials. Alzheimers Dement. Feb 2013;9(1 Suppl):S45–55. doi:10.1016/j.jalz.2011.11.002

26. Ricker JH, Axelrod BN. Analysis of an Oral Paradigm for the Trail Making Test. Assessment. 1994;1(1):47–51. doi:10.1177/1073191194001001007

27. Ricker JH, Axelrod BN, Houtler BD. Clinical validation of the oral trail making test. *Neuropsychiatry, Neuropsychology*, & Behavioral Neurology. 1996;9(1):50–53.

28. Coffin C, Suerken CK, Bateman JR, et al. Vascular and microstructural markers of cognitive pathology. Alzheimers Dement (Amst). 2022;14(1):e12332. doi:10.1002/dad2.12332

29. Duran T, Bateman JR, Williams BJ, et al. Neuroimaging and clinical characteristics of cognitive migration in community-dwelling older adults. Neuroimage Clin. Oct 12 2022;36:103232. doi:10.1016/j.nicl.2022.103232

30. Oishi K, Faria A, Jiang H, et al. Atlas-based whole brain white matter analysis using large deformation diffeomorphic metric mapping: Application to normal elderly and Alzheimer’s disease participants. NeuroImage. 2009;46(2):486–499. doi:10.1016/j.neuroimage.2009.01.002

31. Rolls ET, Huang CC, Lin CP, Feng J, Joliot M. Automated anatomical labelling atlas 3. Neuroimage. Feb 1 2020;206:116189. doi:10.1016/j.neuroimage.2019.116189

32. Dorsey ER, Kluger B, Lipset CH. The New Normal in Clinical Trials: Decentralized Studies. Annals of Neurology. 2020;88(5):863–866. doi:10.1002/ana.25892

33. Udeh-Momoh CT, Jager-Loots CA, Price G, Middleton LT. Transition from physical to virtual visit format for a longitudinal brain aging study, in response to the Covid-19 pandemic. Operationalizing adaptive methods and challenges. Alzheimer’s & Dementia: Translational Research & Clinical Interventions. 2020;6(1)doi:10.1002/trc2.12055

34. 34. Biddle KD, d’Oleire Uquillas F, Jacobs HIL, et al. Social Engagement and Amyloid-β-Related Cognitive Decline in Cognitively Normal Older Adults. The American Journal of Geriatric Psychiatry. 2019/11/01/ 2019;27(11):1247–1256. doi:https://doi.org/10.1016/j.jagp.2019.05.005

35. Sperling RA, Donohue MC, Raman R, et al. Association of Factors With Elevated Amyloid Burden in Clinically Normal Older Individuals. JAMA Neurology. 2020;77(6):735. doi:10.1001/jamaneurol.2020.0387

36. Alegret M, Espinosa A, Ortega G, et al. From Face-to-Face to Home-to-Home: Validity of a Teleneuropsychological Battery. J Alzheimers Dis. 2021;81(4):1541–1553. doi:10.3233/jad-201389

37. Geddes MR, O’Connell ME, Fisk JD, Gauthier S, Camicioli R, Ismail Z. Remote cognitive and behavioral assessment: Report of the Alzheimer Society of Canada Task Force on dementia care best practices for COVID-19. Alzheimer’s & Dementia: Diagnosis, Assessment & Disease Monitoring. 2020;12(1)doi:10.1002/dad2.12111

38. Brearly TW, Shura RD, Martindale SL, et al. Neuropsychological Test Administration by Videoconference: A Systematic Review and Meta-Analysis. Neuropsychology Review. 2017;27(2):174–186. doi:10.1007/s11065-017-9349-1

39. Wadsworth HE, Dhima K, Womack KB, et al. Validity of Teleneuropsychological Assessment in Older Patients with Cognitive Disorders. Arch Clin Neuropsychol. Dec 1 2018;33(8):1040–1045. doi:10.1093/arclin/acx140

40. Seghezzo G, Van Hoecke Y, James L, et al. Feasibility study of assessing the Preclinical Alzheimer Cognitive Composite (PACC) score via videoconferencing. Journal of Neurology. 2021;268(6):2228–2237. doi:10.1007/s00415-021-10403-1

41. Rattanabannakit C, Risacher SL, Gao S, et al. The Cognitive Change Index as a Measure of Self and Informant Perception of Cognitive Decline: Relation to Neuropsychological Tests. J Alzheimers Dis. 2016;51(4):1145–55. doi:10.3233/jad-150729

42. Kaufer DI, Cummings JL, Ketchel P, et al. Validation of the NPI-Q, a brief clinical form of the Neuropsychiatric Inventory. J Neuropsychiatry Clin Neurosci. Spring 2000;12(2):233–9. doi:10.1176/jnp.12.2.233

43. Ismail Z, Agüera-Ortiz L, Brodaty H, et al. The Mild Behavioral Impairment Checklist (MBI-C): A Rating Scale for Neuropsychiatric Symptoms in Pre-Dementia Populations. J Alzheimers Dis. 2017;56(3):929–938. doi:10.3233/jad-160979

